# PEQI: A population dietary quality index for developed countries

**DOI:** 10.1101/2025.02.25.25322860

**Authors:** Konstantinos Christopoulos, Christina Christou, Konstantinos Eleftheriou, Christis Hassapis

## Abstract

**Background:** While there are numerous dietary quality indices for individuals, a longitudinal population-level index is missing from the literature. This article presents a novel population-level dietary index, the Population Eating Quality Index (PEQI) that measures a country’s annual dietary quality.

**Methods:** Using data from the Food and Agriculture Organization and the Global Dietary Database, PEQI comprises of ten waste-adjusted food items for which weighted scores are assigned according to their effect on human health. Associations between the PEQI and health outcomes for a panel of developed countries were studied to further validate the instrument.

**Results:** The index shows good predictive ability regarding life expectancy at birth and premature mortality for an average developed country.

**Conclusions:** PEQI has multiple potential applications in the ecological study of health and nutrition as an exposure or even as an outcome.

## 1 Introduction

1 A dietary pattern (or index) is more likely to capture the effects of diet and food interactions on health, than studying certain foods or food groups (Tapsell et al., 2016). While there are numerous *a priori* dietary quality indices, such as the Diet Quality Index (Patterson et al., 1994), the Health Eating Index (Kennedy et al., 1995; Guenther et al., 2013), the Healthy Diet Indicator (Huijbregts et al., 1997; Berentzen et al., 2013), the Mediterranean Diet Score (Trichopoulou et al., 2003) and its variations (Buckland et al., 2009; Yang et al., 2014), the Dietary Approaches to Stop Hypertension (DASH diet) (Sacks et al., 1995), the Dietary Diversity Score (Kant et al., 1993), the Quantitative Index for Dietary Diversity (Katanoda et al., 2006), and the Dietary Inflammatory Index (Shivappa et al., 2014), among others, they all apply to the individual level.

To the best of our knowledge, two global country-level indices exist to date regarding nutrition. The first attempt was the Global Nutrition Index (GNI) (Rosenbloom et al., 2008). This index was based on three dimensions, namely, nutritional deficiency, obesity prevalence, and food security. Nevertheless, the proxies used for measuring these dimensions raise serious concerns and the index does consider any dietary intake data. The most recent attempt was the International Diet-Health Index (IDHI) (Wang et al., 2020). IDHI is rather complex. It is based on the impact of 11 foods items on 12 cardiometabolic diseases, mediation effects through blood pressure and BMI, as well as disease prevalence, for each age- and sex-specific group in each country. It is therefore also a composite index.

This article introduces the Population Eating Quality Index (PEQI), a novel dietary quality index that measures the quality of nutrition at the country-level. The index allows the comparison of dietary quality between countries on a yearly frequency. It combines data from two sources, namely, the Food Balance Sheets (FBS) of the Food and Agriculture Organization (FAO) of the United Nations and the Global Dietary Database (GDD) to provide a standardized time-series score for each country in the sample.

The PEQI is based on the weighted scores of ten food components. The components represent the major food consumption groups (see Section 2.2) with the addition of alcohol and added sugars (Micha et al., 2015). The component selection was influenced by the Mediterranean diet—for which several studies have shown a positive association with health outcomes and longevity (Eleftheriou et al., 2018; Martinez-Lacoba et al., 2018; Soltani et al., 2019)—as well as from the recommendations of the EAT-Lancet commission (Willett et al., 2019) and the World Health Organization (WHO, 2020).

An exposition of the data and methods including the data sources, PEQI components, waste adjustment, and formulas, can be found in the next section. Section 3 studies the association of the PEQI with health outcomes for the validation of the instrument. The last section concludes the paper.

## 2 Material & methods

### 2.1 Data sources

As mentioned, the PEQI components come from two data sources, the FBS of the FAO (2023) and the Global Dietary Database (GDD, 2022). The rationale behind the use of items from the GDD is just unavailability in the FBS.

The FBS data show a country’s food supply available for human consumption (FA) in a given period (year). These FAO estimates are based on the following elimination methodology: exports, livestock food, foods for manufacture, foods for agriculture, and losses during storage and transportation are subtracted from, and imports are added to the annual production. For the construction of PEQI, the above values refer to kilograms (kg) per capita per year.

The GDD data concern country-level estimates of grams (g) per day (for whole grains and processed meats), or percentage of total kilocalories (kcal) per day (for added sugars). GDD data come from a variety of survey sources and they were not preferred in the case of items available from the FBS due to the frequency of measurement. Moreover, to address the issue of missing values, one has to employ one or several imputation methods. For the construction of the PEQI, we used Last Observation Carried Forward followed by Last Observation Carried Backward to both GDD and FBS components—the rationale being that food item consumption is very slowly time-varying.

### 2.2 Components

The PEQI values are based on ten food items; six beneficial and four detrimental to human health. A summary of the components is given in Table 1.

**Table 1.** Components and weights of the PEQI.

| *Components* | $w^{as}$ | $w^{acm}$ | $w^{ec}$ | *Source* |
| --- | --- | --- | --- | --- |
| *Fruit (total)* | 1.7 | 1.06 | 1 | FBS |
| *Vegetables (total)* | 2 | 1.06 | 1 | FBS |
| *Pulses (total)* | 1.8 | 1.11 | 1 | FBS |
| *Tree nuts (total)* | 1.5 | 1.18 | 1 | FBS |
| *Whole grains (total)* | 1.6 | 1.27 | 1 | GDD |
| *Fish and seafood (total)* | 1.3 | 1.08 | 1 | FBS |
| *Processed meat (total)* | -1.8 | -1.28 | -1 | GDD |
| *Red meat (bovine, pig, mutton and goat)* | -1.4 | -1.16 | -1 | FBS |
| *Added sugars (total)* | -1.5 | -1.07 | -1 | GDD |
| *Alcoholic beverages (total)* | -2 | -1.52 | -1 | FBS |

Notes: Source refers to the databases; FBS=Food Balance Sheets, GDD=Global Dietary Database. $w$ refers to alternative weights for the final weighting in the PEQI formula in Equation 2; see Section 2.4.2 for details.

The six beneficial foods are fruits, vegetables, pulses, tree nuts, whole grains, and fish and seafood. The benefits of fruit and vegetable consumption are well established. Specifically, increased consumption is associated with decreased risk for all-cause mortality and cardiovascular mortality in particular (Wang et al., 2014). Pulses—according to FAO, refer to the the dry seeds of non-oilseed legumes such as beans, broad beans, peas, chickpeas, cowpeas, pigeon peas, lentils, bambara beans, vetches, lupins, and other “minor” pulses—have been linked with reduced cardiovascular disease (CVD) and coronary heart disease (CHD) incidence, as well as with lower hypertension, and obesity incidence (Viguiliouk et al., 2019). Tree nuts—refer to walnuts, pistachios, macadamia nuts, pecans, cashews, almonds, hazelnuts, and Brazil nuts—have been associated with cardioprotective benefits, as well as with decreased cancer and respiratory disease risk (Balakrishna et al., 2022). Whole grain consumption has been linked with reduced risk for CVD, cancer, and all-cause mortality, as well as with lower mortality from respiratory and infectious disease, and diabetes (Aune et al., 2016). Fish and seafood consumption, despite potential contamination, appears to benefit a variety of atopic, musculoskeletal, gastrointestinal, and ophthalmologic outcomes. It is also associated with a slight reduction in the risk of several CVDs, as well as of gastrointestinal cancer, metabolic syndrome, dementia, and Alzheimer’s disease (Li et al., 2020).

The four chosen food items detrimental to human health are processed meat, red meat, added sugars, and alcohol. Processed meat consumption is associated with a variety of cancers (prostate, breast, colorectal, pancreatic) as well as with CVD mortality and diabetes (Wolk, 2017). Red meat (mainly beef and pork, but also lamb, venison, boar, mutton, and goat) consumption is mainly associated with increased risk for colorectal and advanced prostate cancer as well as with CVD mortality (Wolk, 2017). Sugars that have been added to foods during processing or preparation (not occurring naturally) are considered to be added sugars. While there is no scientific consensus on whether the added sugars *per se* play a role in obesity, diabetes, or CVD (Rippe and Angelopoulos, 2016), foods containing them probably present a health hazard. Therefore, this component also serves as a proxy for ultra-processed ‘junk’ food consumption. Lastly, alcohol consumption is causally related to numerous medical conditions and presents one of the largest burdens of disease globally (Room et al., 2005). Aside from liver damage, heavy alcohol consumption may lead to a variety of gastrointestinal, but not only, cancers. Much evidence exists also for mental health disorders and CVD (Rehm et al., 2010).

### 2.3 Waste adjustment for FBS components

All components from the FBS were waste adjusted following the FAO methodology (Gustavsson et al., 2011) as was adapted by the World Health Organization (WHO, 2023), so that values approximate actual food consumption and not mere availability. The level of consumption is given by the following general formula:

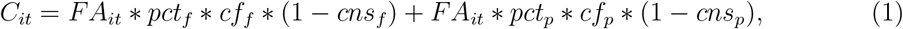

where *C*_*it*_ and *FA*_*it*_ are the consumption and the food availability for country *i* at year *t*, respectively. *cf*_*f*_ and *cf*_*p*_ are the conversion factors that convert the total volume to the edible part for the fresh and processed form, respectively. The conversion factors used are given in Table 2. *pct*_*f*_ and *pct*_*p*_ are the proportions used as fresh and processed (*pct*_*f*_ = 1 *− pct*_*p*_), respectively. *cns*_*f*_ and *cns*_*p*_ are the respective percentages of fresh and processed food wasted during consumption. All proportions are in decimal form.

**Table 2.** Conversion factors for PEQI components from FBS.

| *Components* | $cf_f$ | $cf_p$ |
| --- | --- | --- |
| *Fruits* | 0.8 | 0.75 |
| *Vegetables* | 0.8 | 0.75 |
| *Pulses* | 0.79 | 0.79‡ |
| *Tree nuts* | 0.79 | 0.79‡ |
| *Fish and seafood* | 0.5 | 0.5† |
| *Beef* | 0.715 | 0.715‡ |
| *Pork* | 0.68 | 0.68‡ |
| *Other meat* | 0.7 | 0.7‡ |

Notes: Conversion factors for beef, pork, and other meat are used for the waste adjustment of the respective red meat components, that is, bovine, pig, and mutton and goat meat. The processing of meat here refers to industrial mechanical processing involving cutting, grinding, and mixing, and not processing for flavour and shelf life enhancement. †A common $cf$ for fish and seafood is provided by WHO (2023). ‡A common $cf$ for pulses, tree nuts, and red meat components is implied by WHO (2023).

Factors other than the conversion factors (*cf*) differ not only by component but also by region. These data can be found in Table 3 and in more detail in WHO (2023). The right hand side of Equation 1 has two parts, a ‘fresh’ denoted by the subscript *f* and a ‘processed’ denoted by the subscript *p*, which penalize the respective food availability. The distinction between fresh and processed (i.e., conversion and waste adjustment factors, and proportions used being different between fresh and processed food) applies to fruits, vegetables, and fish and seafood. For the rest of the FBS components, a common *cf, pct*, and *cns* is applied. The component specific formulas for waste adjustment can be found in Table 4.

**Table 3.** Food waste and processing proportions by component and region.

| *Region/Components* | *Fruits & Vegetables* |  |  | *Tree nuts & Pulses* | *Fish & seafood* |  |  | *Red meat* |
| --- | --- | --- | --- | --- | --- | --- | --- | --- |
| *item* | $pct_p$ | $cns_f$ | $cns_p$ | $cns†$ | $pct_p$ | $cns_f$ | $cns_p$ | $cns†$ |
| *Europe* | 0.60 | 0.19 | 0.15 | 0.04 | 0.96 | 0.11 | 0.10 | 0.11 |
| *USA, Canada, Oceania* | 0.60 | 0.28 | 0.10 | 0.04 | 0.96 | 0.33 | 0.10 | 0.11 |
| *Industrialized Asia* | 0.04 | 0.15 | 0.08 | 0.04 | 0.96 | 0.08 | 0.07 | 0.08 |
| *N. Africa, West & Cent. Asia* | 0.50 | 0.12 | 0.01 | 0.02 | 0.96 | 0.04 | 0.02 | 0.08 |
| *Latin America* | 0.50 | 0.10 | 0.01 | 0.02 | 0.96 | 0.04 | 0.02 | 0.06 |

Notes: † $cns_p = cns_f = cns$

**Table 4.** Waste adjustment formulas for the PEQI components from FBS.

| *Components* | *Formula* |
| --- | --- |
| *Fruits* | $FA_{it} * (1 - pct_p) * cf_f * (1 - cns_f) + FA_{it} * pct_p * cf_p * (1 - cns_p)$ |
| *Vegetables* | $FA_{it} * (1 - pct_p) * cf_f * (1 - cns_f) + FA_{it} * pct_p * cf_p * (1 - cns_p)$ |
| *Pulses* | $FA_{it} * cf * (1 - cns)$ |
| *Tree nuts* | $FA_{it} * cf * (1 - cns)$ |
| *Fish and seafood* | $FA_{it} * (1 - pct_p) * cf_f * (1 - cns_f) + FA_{it} * pct_p * cf_p * (1 - cns_p)$ |
| *Bovine meat* | $FA_{it} * cf * (1 - cns)$ |
| *Pig meat* | $FA_{it} * cf * (1 - cns)$ |
| *Mutton and goat meat* | $FA_{it} * cf * (1 - cns)$ |

Notes: Bovine, pig, and mutton and goat meat are the food items that comprise red meat.

### 2.4 Scores, weights, and the PEQI formula

#### 2.4.1 Scores

Nine percentiles, namely p10, p20, p30, p40, p50, p60, p70, p80, and p90, were calculated from the pooled data of countries (see Section 3) for each component and a score from one to ten was assigned in the following manner. For consumption less than p10, a score of one was assigned. For consumption more or equal to p90, a score of 10 was assigned. Accordingly, scores were given for values between the other percentiles. The rationale behind calculating percentiles using the pooled sample—and not each year separately— is that the PEQI should adhere to an *a priori* dietary pattern and enable comparability, both within and between countries.

The use of face values of food consumption (e.g., kg/per capita) from FBS or GDD to determine the PEQI score was deemed inappropriate for two main reasons. First, despite waste adjustment, these ecological data remain population-averaged values with measurement error and comparing them to dietary recommendations (or other evidence) would result in false inference. Second, the majority of the items comes from the FBS where measurement error is expected to be larger. Therefore, percentiles allow a valid calculation of the index despite these issues, under the assumption that the errors are more or less the same for each country.

#### 2.4.2 Weights

For the calculation of the PEQI index, we suggest using the weighted scores introduced in Section 2.4.1. Three different weights were tested (see Table 1); *w*^*ec*^=equal contribution weights, *w*^*as*^=authors’ suggested weights, and *w*^*acm*^=all-cause mortality weights.

*w*^*as*^ weights were chosen based on the strength of evidence regarding cancer and cardiovascular outcomes from the World Cancer Research Fund (WCRF, 2018) and (Brandhorst and Longo, 2019), respectively. Therefore, they give emphasis on the two most pertinent chronic diseases in the developed world. Nevertheless, they remain somewhat subjective and unreproducible.

The construction of *w*^*acm*^ is based on a more objective approach. Specifically, Hazard ratios (HR) from meta-analyses on the effect of food items on all-cause mortality were converted—so that *HR ≥* 1 for symmetry—and used as weights. The HR were taken from Schwingshackl et al. (2017) with the exception of alcohol, where estimates from Jayasekara et al. (2014) were used, due to availability. The chosen HR was the one whose serving approximated 100g per day, in an effort to adjust for consumption. For added sugars, the estimate for the sugar-sweetened beverages was used since these drinks account for the majority of added sugars in a Western diet (Sánchez-Pimienta et al., 2016).

Lastly, despite the fact that not all components are expected to affect human health equally, simple -1, 1 weights (*w*^*ec*^) were also tested.

#### 2.4.3 Formula and discussion

The PEQI for a given country *i* and year *t* is a weighted sum given by the following formula:

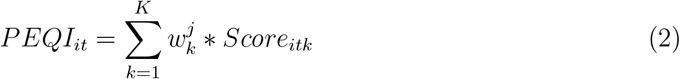

where *w*^*j*^ is the chosen weight *j* (*j* = *w*^*as*^, *w*^*acm*^, *w*^*ec*^) of the component *k* from Table 1. The index was calculated with all three alternative weights. The index is then standardized by subtracting the mean and dividing by the standard deviation of the pooled data of countries (see Section 3). In this way, results are easier to interpret whether the index is used as a covariate or as a dependent variable.

At this point, we have to note that both the selection of components and weights contain a degree of objectivity. This may be more true for the weight selection. Nevertheless, as it is evident from the analysis in Section 3, the index demonstrated insensitivity to weight selection, at least for the weights examined. However, since equal contribution weights (*w*^*ec*^) would constitute an unrealistic assumption, authors’ recommend the use of one of the other weights.

All in all, the PEQI does not preclude alternate weights to be used or calculated, or even estimated. For example, one can find the weights that result into a PEQI by maximizing its association with a health outcome via an iterative process. But this approach has two issues: First, what would this outcome be—given that different outcomes will ‘select’ different weights—and second, this procedure feels a lot like p-hacking.

## 3 Empirical application: Associations with health outcomes

An eating quality index must demonstrate predictive ability in health outcomes, even at the country level. To this end, associations with two health outcomes were examined for a panel of developed countries.^1^ The health outcomes examined were life expectancy at birth and premature mortality. The latter was measured by age-standardized potential years of life lost (PYLL) rates with 75 as the cut-off age. The data for the two health outcomes were extracted from the OECD health database (OECD, 2016).

Bayesian hierarchical models (see Appendix A for the methodological details)—using all three weights (*w*)—were chosen for the empirical analysis due to their regularization properties. The findings were quantitatively similar irrespective of the weight *w* used. The standardized PEQI (PEQI std) was positively associated with life expectancy at birth and negatively associated with premature mortality. More specifically, a one standard deviation increase in the standardized PEQI was associated with 3.36 (0.14), 3.45 (0.15), and 3.23 (0.15) years increase in life expectancy (standard errors are in parentheses) for *w*^*ec*^, *w*^*as*^, and *w*^*acm*^ weights, respectively.

For premature mortality, a one standard deviation increase was associated with a 27%, 28%, and 26% rate reduction in potential years of life lost—standard error was 0.01, in the original scale, for all three weights—for *w*^*ec*^, *w*^*as*^, and *w*^*acm*^, respectively. Figure 1 depicts the posterior retrodictive simulation for the average country for life expectancy and premature mortality using *w*^*as*^. A more detailed exposition of the results can be found in Appendix B.

**Figure 1:**
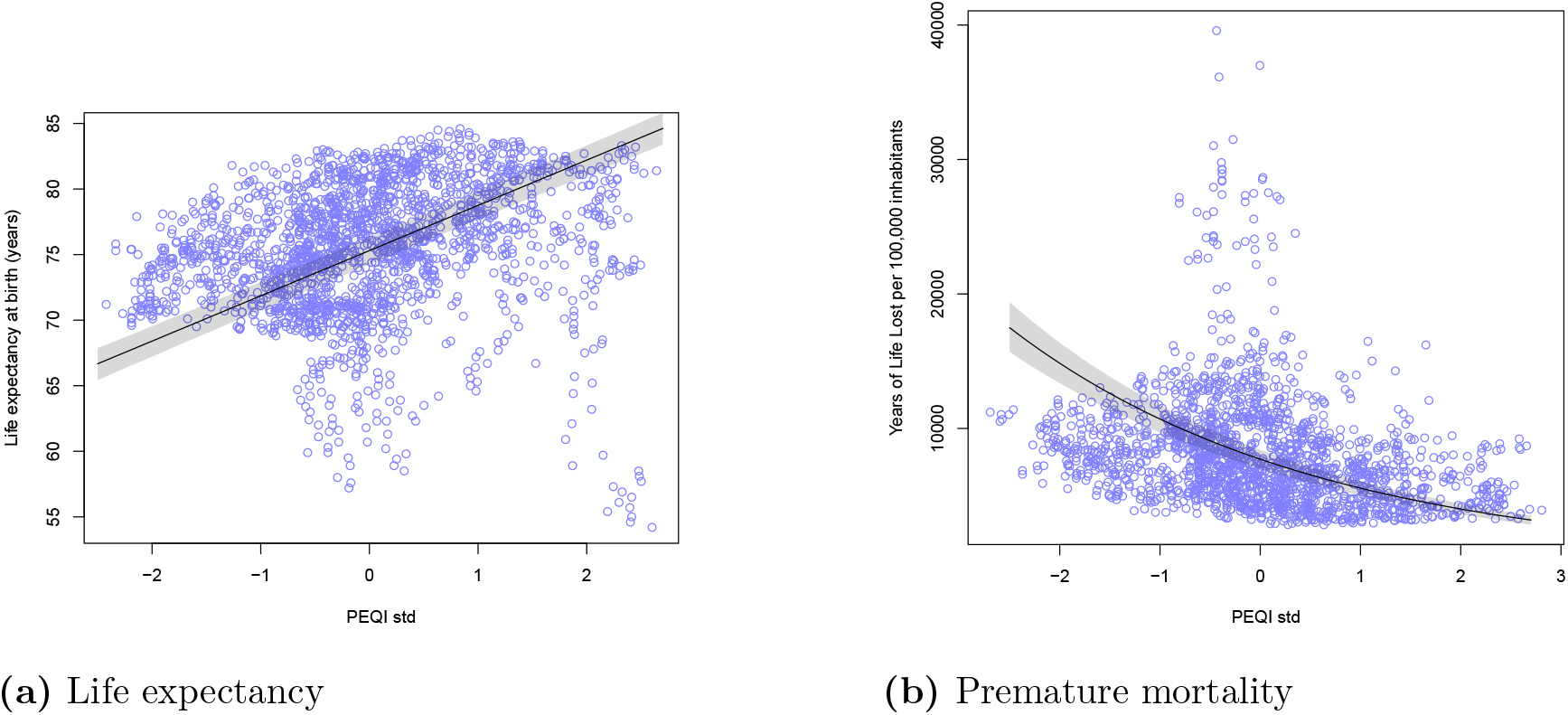
Posterior predictive simulation for life expectancy (a) and premature mortality (b) for an average country. Shaded regions are 89% highest posterior density intervals.

## 4 Conclusions

The PEQI is, to the best of our knowledge, the first instrument developed to measure the dietary quality of a developed country longitudinally in a comparable manner without the inclusion of other factors. It has many potential applications in studies of health and nutrition but also in studies where nutritional quality has to be adjusted for.

This index was developed mainly for developed countries. This methodology is probably not adequate to capture dietary quality in a developing country with a severely malnourished population, food shortages, and minimal processing of foods. Nor is it recommended to compare countries close to the extremes. The development of an index for developing countries constitutes a topic for future research. The index is also sample dependent inherently, that is, a different panel of countries and years will result in different scores. It is therefore important to specify if the values were drawn from this paper or calculated *de novo*.

In sum, this article presents the PEQI, a novel dietary index for cross-country comparison of nutritional quality. The index adjusts for wasted food before the calculation of its scores. Its validity is demonstrated by strong associations with increased life expectancy and decreased premature mortality. It is not sensitive to weight selection and its standardization is strongly recommended for its use in models.

## Data Availability

All data produced in the present study are available upon reasonable request to the authors

## Declarations

### Ethics approval and consent to participate

Not applicable

### Consent for publication

Not applicable

### Availability of data and materials

The data and code used for the analysis will be made available on reasonable request.

### Authors’ contributions

K.C. and K.E. conceptualized and designed the study. K.C. led the data analysis, interpretation, and wrote the manuscript. C.C., K.E., and C.H. contributed to the writing.

## Acknowledgements

Authors would like to thank Marco Springmann for helpful suggestions regarding food data not available from FAO.

## Appendix A: Statistical analysis

For the analysis, Bayesian hierarchical models with random intercepts and weakly informative priors were employed (see Chapter 5 in Gelman et al. (2013) for an introduction). These models allow for the partial pooling of information across clusters using an adaptive regularizing prior which results in shrinkage toward the mean, and therefore, better estimates for the between cluster differences, especially in imbalanced sampling (McElreath, 2018).

For life expectancy (LE) the following linear model was used

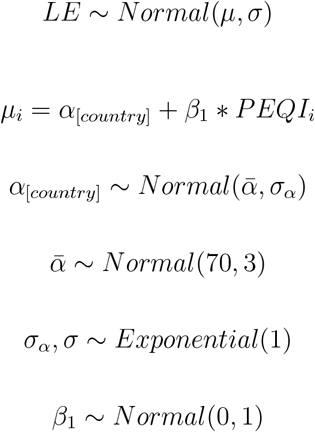

where *α*_[*country*]_ are the random country intercepts, *i* denotes the observation, and PEQI the standardized 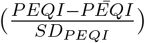 index. For premature mortality (PM) the following Gamma-Poisson model was used

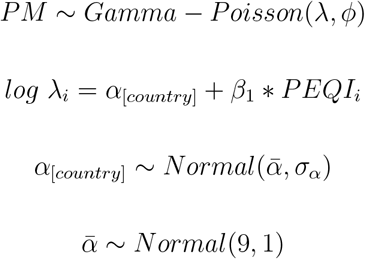

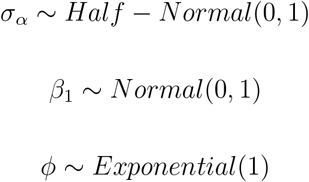

The analysis was performed using Hamiltonian Monte Carlo sampling as implemented in the *R* package *‘rethinking’* (McElreath, 2018).

## Appendix B: Results

Forest plots and posterior density graphs with 89% highest posterior density intervals (HPDI) for each health outcome and each weight, *w*, are depicted in Figure 2. The first column of Figure 2 refers to life expectancy and the second column to premature mortality. The black, blue, and red curves are the posterior densities of *β*_1_ (exponentiated for the premature mortality models) for *w*^*as*^, *w*^*acm*^, and *w*^*ec*^, respectively.

**Figure 2:**
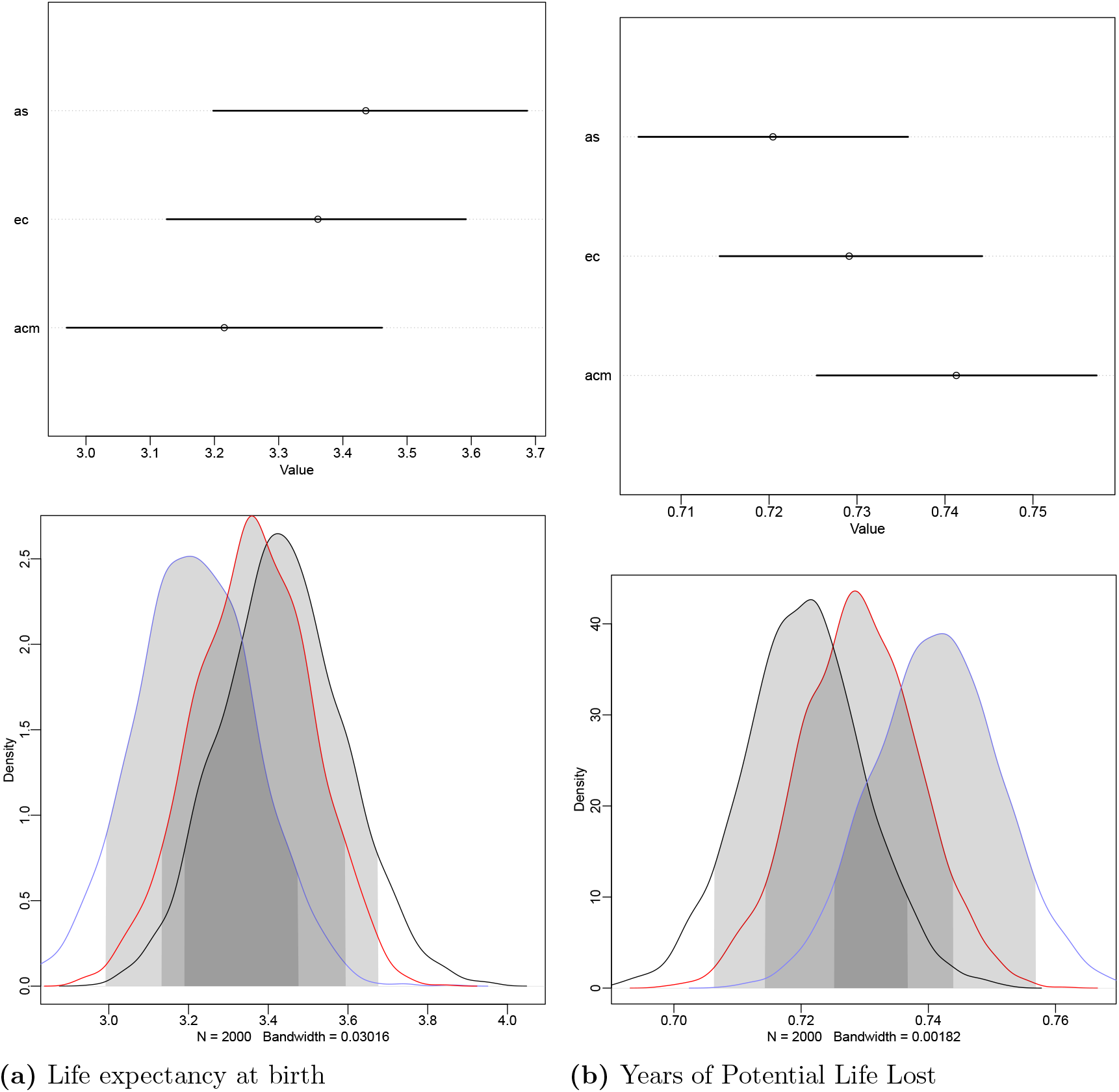
Forest plots (first row) and posterior densities (second row) with 89% HPDI of the association of PEQI with life expectancy (a) and premature mortality (b). For forest plots *as, acm*, and *ec* correspond to the respective weights. *w*^*as*^ (black), *w*^*acm*^ (blue), and *w*^*ec*^ (red).

## Footnotes

1 Australia, Austria, Belgium, Bulgaria, Brazil, Canada, Switzerland, Chile, Colombia, Costa Rica, Czech Republic, Germany, Denmark, Spain, Estonia, Finland, France, Great Britain, Greece, Croatia, Hungary, Ireland, Iceland, Isreal, Italy, Japan, South Korea, Lithuania, Luxembourg, Latvia, Mexico, Netherlands, Norway, New Zealand, Poland, Portugal, Romania, Slovakia, Slovenia, Sweden, Turkey, and the United States. The years span from 1970 to 2021.

